# Deep Time-to-Event Models for Intrapartum Fetal Monitoring

**DOI:** 10.64898/2026.09.21.26363416

**Authors:** Guilherme S. Imai Aldeia, Helena Coggan, Yuting Yang, Lisa Levine, Jennifer A. McCoy, William G. La Cava

## Abstract

Timely detection of fetal distress during labor is a central pursuit of modern obstetric care. The primary measure used in this regard is fetal heart rate and contraction monitoring (echocar-diotocography or CTG), which provides a continuous signal during labor. The vast majority of machine learning applications to CTG assume access to a 30 to 60 minute window just prior to delivery, which makes an untenable assumption that the timing of delivery is known. In this work, we reframe the problem as a time-to-event prediction task. We develop deep time-to-event models to predict the joint probability of imminent delivery and fetal acidosis, a biomarker of fetal hypoxia collected from umbilical cord gas after delivery. We develop a framework for this problem, drawing on deep discrete-time survival methods and marked point processes, adapting each to this particular setting (i.e., continuous physiological signals, fully observed events, partially observed labels). One approach dubbed Marked DeepHit accurately estimates *acidemic delivery* within one hour at multiple elapsed-time land-marks, with test-set AUROC of 0.837 [95% CI 0.692–0.970] and 0.935 [0.858–0.964] at 6 and 12 hours into labor, respectively. We find that these models also achieve competitive performance near delivery time compared to prior models trained specifically on the end of tracing, and outperform other clinical feature-based proposals, both on internal and external validation at a second site. These results give a proof-of-principle to support prospective evaluation of AI-EFM during labor.

**Data and Code Availability:** We use data from a multi-site academic health system (University of Pennsylvania Health System) and the CTU-UHB repository (Chudáček et al., 2014). Access to CTU-UHB is openly available under the terms of the data use agreement on Physionet. Code for this paper is available at https://github.com/cavalab/deep-tte-intrapartum-fetal-monitoring.

**Institutional Review Board (IRB):** This study was approved by the Institutional Review Board at University of Pennsylvania under a waiver of informed consent.

## 1. Introduction

Continuous electronic fetal monitoring (EFM) records fetal heart rate (FHR) and uterine activity (tocodynamometry, TOCO) throughout most U.S. labors (Martin et al., 2003). Despite its ubiquity, continuous CTG has not clearly reduced perinatal mortality or cerebral palsy, and inter-rater agreement on categorical interpretations is modest (Alfirevic et al., 2017; Nelson et al., 1996; Macones et al., 2008), motivating automated interpretation (Georgieva et al., 2019).

Arterial cord pH is an objective, routinely collected surrogate of intrapartum acid-base status associated with neonatal morbidity and longer-term neurologic outcomes (Malin et al., 2010). Deep learning applied to EFM (AI-EFM) has predominantly used *terminal classifiers* : given a tracing window near the end of labor, predict whether pH falls below a threshold (Spilka et al., 2017; Petrozziello et al., 2018; McCoy et al., 2024).

AI-EFM is intended for clinical decision support throughout labor, but terminal classifiers use retrospectively-selected end-of-labor segments. Labor timing varies substantially: the median time from painful contractions to active labor was 16.0 [10.0–26.6] hours in nulliparous patients and 9.4 [5.9– 15.3] hours in multiparous patients in a large cohort study (Tilden et al., 2023). Timing is also difficult to predict: models using maternal characteristics with or without cervical dilation had AUROC 0.65–0.66 for spontaneous labor onset (Sanusi et al., 2023), consistent with the lack of a clinically accepted objective measure of cervical change (Feltovich, 2017). Thus, knowing that a tracing is near delivery is itself a difficult prerequisite for deploying terminal classifiers. An acidemic delivery in 20 minutes and one in 14 hours have the same label but very different clinical implications.

Terminal classification misses the important element of time. We therefore formulate EFM-based acidemia prediction as a *landmark time-to-event* problem. We define the target as acidemic delivery within a horizon, rather than acidemia conditional on imminent delivery. We propose competing-risks and marked point-process models (Fine and Gray, 1999; Lee et al., 2018; Du et al., 2016; Shchur et al., 2021) for a terminal delivery event with an ordinal, partially observed acidemia mark. Both retain delivery-time supervision when cord pH is missing, enabling training on more than 100,000 labor tracings (1.2 million monitoring hours), and both are evaluated at elapsed- and end-time landmarks.

## 2. Background

### 2.1. Computerized EFM Interpretation

Computerized CTG systems have long extracted expert-derived EFM features, with clinical evidence reviewed extensively (Ben M’Barek et al., 2023; Mendis et al., 2023; Campanile et al., 2020); examples include the fetal reserve index or the final-120-minute deceleration area (AUROC 0.76 [0.72–0.80] for acidemia) (Evans et al., 2023; Cahill et al., 2018). AI-EFM studies use convolutional networks, LSTMs, transformers, and related models (Petrozziello et al., 2018, 2019; McCoy et al., 2024; Mendis et al., 2025), typically framing fetal compromise as a binary outcome in a fixed end-of-tracing segment. Many use CTU-UHB, which contains 522 final-90-minute tracings (Chudáček et al., 2014). Prior work applied a classifier within the latest 60 minutes of CTU-UHB recordings (Mendis et al., 2024), and has modeled acidemic-delivery hazards in the second stage using clinician-assigned CTG categories and obstetric covariates (Cavoretto et al., 2023).

Existing terminal models do not establish prospective, time-indexed risk. No prior AI-EFM study has estimated the horizon-specific joint probability of delivery and acidemia from an ongoing tracing. Here we distinguish imminent acidemic delivery from later acidemia while estimating time remaining to delivery.

### 2.2. Time to Event Models

Time-to-event models estimate the probability of an event within any chosen horizon. With *competing risks*, the cumulative incidence gives the probability of a particular mutually exclusive event type by that horizon (Cox, 1972; Fine and Gray, 1999; Andersen et al., 2012; Putter et al., 2007). DeepHit directly learns discrete-time cumulative incidence functions (Lee et al., 2018). *Marked point processes* model events and their associated labels (Du et al., 2016; Reinhart, 2018; Shchur et al., 2021). Our marked model factorizes the joint probability into delivery timing and acidemia. *Landmarking* applies the model repeatedly as information arrives, conditioning on the history available at each landmark and predicting residual time without future observations (van Houwelingen, 2007; Zheng and Heagerty, 2005). For EFM, delivery is the terminal event and cord pH is its mark.

## 3. Methods

### 3.1. Task Formulation

For patient *i* we observe a multichannel physiological signal *X*_*i*_(*u*)∈ ℝ^*d*^ sampled over an interval [0, *τ*_*i*_], where *u* is time since the start of the retained tracing and *τ*_*i*_ is the final tracing sample, taken to coincide with delivery. The channels are FHR and TOCO.

Let *M*_*i*_ *∈* ℝ denote the cord pH at delivery, and let *R*_*i*_ = 1 when it is observed and *R*_*i*_ = 0 otherwise. For a threshold *c* define the acidemia indicator:

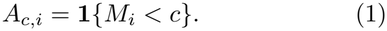

Thus, *A*_*c,i*_ is observed only when *R*_*i*_ = 1.

Prediction is made at a *landmark* time *t ∈* [0, *τ*_*i*_). Let *X*_[*a,b*]_ ={ *X*(*u*) : *a ≤ u ≤ b}* be the segment of tracing between two times. At the landmark, the only admissible information is *X*_[0,*t*]_, namely everything recorded up to *t*. The quantity of interest is the *remaining* time to delivery

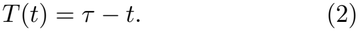

We drop the patient subscript *i* except where pairs of patients are compared. For a fixed landmark, we abbreviate *T* = *T* (*t*). In this study, a model does not ingest all of *X*_[0,*t*]_ but a fixed-width window *x* = *X*_[*t−w, t*]_ of width *w* ending at the landmark. In principle the model could ingest a sequence of such windows recurrently, which we do not explore here.

This is referred to as the landmarking construction of dynamic prediction (van Houwelingen, 2007; Zheng and Heagerty, 2005): rather than modeling the full longitudinal process jointly with the event, we fit a model of the residual event time conditional on *X*_[0,*t*]_, and re-apply it as *t* advances.

### 3.2. Estimands

The main clinical target is the probability that an *acidemic delivery* occurs within a forecast horizon *h*:

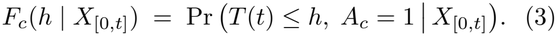

This joint risk factors into imminent-delivery risk and conditional pH risk:

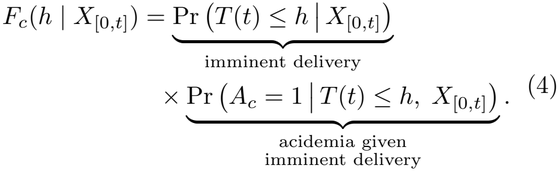

Terminal classifiers for EFM (Spilka et al., 2017; Cahill et al., 2018; Petrozziello et al., 2019; McCoy et al., 2024) estimate only the second term, i.e., the severity of pH *given* that delivery is imminent:

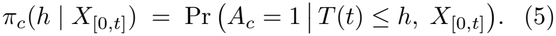

The conditional-pH-risk estimand in Eq. (5) conditions away the delivery-timing question entirely. It is appropriate for decisions when delivery is known, but cannot alone define an alert criterion.

We report the probability of delivery within the time horizon, Pr(*T* (*t*) *≤ h* |*X*_[0,*t*]_), as a third target, both because it is clinically useful and because it is the factor that separates the joint-risk estimand in Eq. (3) from the conditional-pH-risk estimand in Eq. (5). Forecasting delivery timing distinguishes an imminent acidemic delivery from an acidemic delivery that is still remote, and determines whether a pH-risk prediction can support a time-sensitive intervention.

### 3.3. Deep Time to Event Models

Our problem differs substantially from that of conventional survival analysis: delivery is an observed terminal event for every retained pregnancy, whereas cord pH is an ordinal mark missing for some deliveries. Thus, missing pH denotes an unobserved delivery mark, not a censored delivery: the delivery is observed for every retained pregnancy, and its timing remains known. As a result, the standard discrete-time survival model does not fit neatly onto this task. Following DeepHit (Lee et al., 2018) as implemented in `pycox` (Kvamme et al., 2019), we discretize remaining time into *L* bins with upper boundaries *κ*_1_ *< … < κ*_*L*_ and write *l*(*s*) *∈ {*1, …, *L*} for the bin containing duration *s*. The final layer produces logits *ϕ∈* ℝ^*J×L*^, where *J* indexes “causes” (e.g., an acidemic outcome category) and *J* = 1 in the single-risk case.

The challenge is therefore to design a loss function that can be optimized effectively, encourages the outputs to form a valid cumulative probability distribution, and allows the model to leverage the temporal structure of the problem. We consider two parameterizations of this delivery process: one directly models the joint distribution of delivery time and pH category; the other factors these into delivery-time and delivery-time-conditioned pH-mark distributions. For clarity and brevity some equations in this section are shown in Appendix A.

#### 3.3.1 Competing risks: a joint cause–time model

For the competing-risks formulation, observed pH values are partitioned into mutually exclusive categories defined by different thresholds, ordered from most to least acidemic. Let *E*_*j*_ *∈ {*1, …, *J}* denote TTE Models for Fetal Monitoring this category, so that *A*_*c*_*k,i* = **1 {***E*_*j*_ *≤ k}* . The joint cause–time model directly parameterizes the probability mass of each delivery-time and pH-category combination:

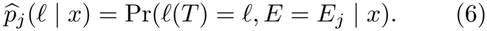

We define the cause-specific cumulative incidence as 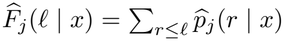. Risk at a pH category *c*_*k*_ is then a partial sum over categories, calculated as:

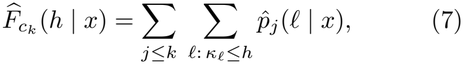

This cumulative incidence function is automatically monotone in *k*, allowing one model to serve every threshold.

Following competing-risks DeepHit (Lee et al., 2018), pH-observed rows contribute a convex combination of a likelihood term and a ranking term. Eqs. A.2–A.5 give the full partial-label loss. We extend this objective to pH-missing deliveries by marginalizing their likelihood over pH categories. Each batch is split into pH-observed samples, *O*, and pH-missing samples, ℳ. The compact batch objective is:

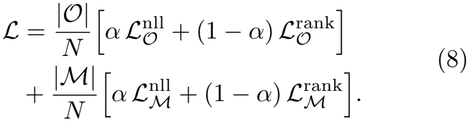

Here, *α* ∈ [0, 1] trades off the observed-cause likelihood and ranking terms. For pH-missing samples, *α* and (1 − *α*) weight the marginal likelihood and all-cause ranking loss, respectively. The cause-specific and all-cause ranking terms are evaluated within *O* and ℳ, respectively, so each sample contributes to one likelihood and one ranking objective.

#### 3.3.2. Marked point process: a factored time–mark model

As in temporal marked point processes (Reinhart, 2018), the second formulation we propose factors the joint distribution of delivery time (the event) and pH (the mark) into a timing distribution and a time-conditioned mark distribution:

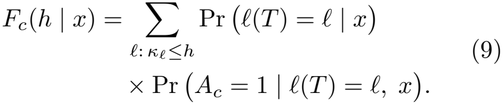

The two factors are modeled by distinct heads that make use of a shared encoder. The delivery head emits logits *ϕ∈* ℝ^*L*^ over time bins, converted to 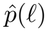 by the discrete-time softmax in Eq. (A.1) with *J* = 1. For the marked formulation, *K* = 4 denotes the number of pH thresholds. The mark head emits logits *ψ ∈* ℝ^*L×K*^ —one per time bin and threshold—with:

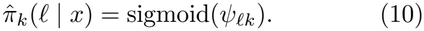

This is the conditional probability that pH falls below *c*_*k*_ given delivery in bin *l*. The marked time– risk factorization in Eq. (9) gives the predicted risk,

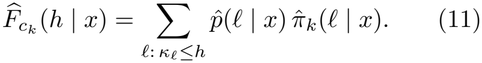

The model’s conditional pH risk, corresponding to the conditional-pH-risk estimand in Eq. (5), is 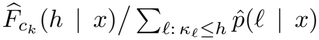 whenever the denominator is positive.

Only rows with observed pH supply a mark target. They define the mark loss, ℒ^mark^, a binary cross-entropy over thresholds evaluated at the *observed* delivery bin and masked by *R*_*i*_ (Eq. A.6).

Let 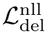 and 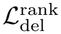 denote the ordinary single-risk DeepHit likelihood and ranking losses, respectively, evaluated on all *N* rows. Then the full objective is:

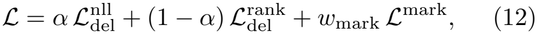

#### 3.3.3. Formulation comparison

The competing risks and marked point process formulations have practical differences worth noting. Both can train on all labors, including those without pH results; the marked model’s delivery head does this with the ordinary single-risk objective, whereas the competing-risks model requires a partial-label objective (Eq. (A.4)). These are subtly different targets, as the competing risks training specifically pressures correct delivery time ranking within pH categories when pH labels are present. Second, the marked model’s *K* binary mark heads are trained independently, so threshold nesting is not guaranteed under the binary cross-entropy objective; the categorical competing model gets nesting for free from the threshold cumulative-incidence calculation in Eq. (7) but must fit rare categories directly. Third, both models can obtain the conditional-pH-risk estimand in Eq. (5) by dividing their threshold-specific cumulative incidence by all-cause delivery risk; however, the marked model differs by explicitly parameterizing the time-conditioned pH-mark probabilities that enter this ratio.

### 3.4. Data Curation

The development set comprised intrapartum EFM from the multi-site University of Pennsylvania Health System (2006–2020). Most sites have a universal cord-gas collection policy, mitigating the selective testing common elsewhere (Kearney et al., 2024).

We linked CTG tracings to arterial cord-gas pH by patient identifier, retaining lab results timestamped within 30 minutes of EFM end and excluding possible multiple pregnancies. We used pH thresholds of 7.2, 7.15, 7.1, and 7.05.

#### Signal Processing

CTG was resampled at 0.25 Hz, capped at 48 hours, and modeled in one-hour (900-sample) windows. We retained windows with at most 30% raw input-channel missingness before imputation. During training, inputs were forward-filled with observation-mask and elapsed-time channels.

### 3.5. Model Architectures and Training

Both formulations use a 128-dimensional Inception-Time encoder (Ismail Fawaz et al., 2019, 2020), selected because it performed best in prior EFM work (McCoy et al., 2024). Its modules combine bottlenecked parallel convolutions and max pooling with residual connections and global average pooling.

Models output 11 discrete delivery-time bins, corresponding to 0, 10min, 20min, 40min, 1h, 2h, 4h, 8h, 14h, 24h, and 48h. The competing-risks head consists of five independent cause-specific MLPs, each producing logits for the 11 delivery-time bins. Their outputs are stacked into a 5*×* 11 cause–time logit array and jointly normalized by the competing-risks softmax.

We used fixed train, validation and test splits with no overlapping patients. Validation was used for model selection, early stopping, and calibration. Test data were reserved for final evaluation. Training used Adafactor with cosine annealing (Loshchilov and Hutter, 2017), up to 100 epochs, 30-epoch early stopping, and returned the lowest-validation-loss checkpoint.

To ensure calibrated probabilities, Platt scaling was applied after training at each discrete prediction horizon, including a parameter for elapsed time, using the validation set.

## 4 Experiments

We evaluated marked point-process and competing-risks models for delivery timing and pH-threshold marks from FHR/TOCO tracings and post-delivery laboratory measurements. Appendices B and C report architecture and sampling strategy.

### 4.1. Metrics

At each landmark, evaluation used only information available up to that time. We evaluated start-landmarks at 1, 6, 12, and 20 hours with 1-, 2-, and 4-hour horizons, and end-landmarks at 0, 20, 40, and 60 minutes before delivery. The start-landmarks were clinically interpretable whole numbers that approximately divide observed tracing lengths into quartiles; the 1-, 2-, and 4-hour horizons were selected with maternal-fetal medicine clinicians as actionable review windows. End-landmarks permit comparison to end-of-tracing classifiers. Our primary measure was *acidemic delivery by horizon*: AUROC for acidemic delivery within a horizon, with all other patients as controls. Horizon-specific AUROC and Antolini’s time-dependent concordance both assess ranking but answer different questions: the former distinguishes an imminent acidemic delivery from outcomes outside the chosen window, whereas the latter aggregates comparable-pair rankings across observed event times. We used 100 patient-level percentile-bootstrap resamples for 95% confidence intervals.

### 4.2. Baselines

We evaluated three baselines. We used a transparent CTG-only, five-component approximation of the Fetal Reserve Index with Cox regression for acidemic delivery, called *Rule Based*. It scores FHR baseline in the FIGO reference range (110–160 bpm), FHR variability in range (5–25 bpm), an acceleration, fewer than three FIGO-defined decelerations per 30 minutes, and no tachysystole (Evans et al., 2023; Ayres-de-Campos et al., 2015). We trained landmark-specific *XGBoost* Cox models using 23 EFM features from Vargas-Calixto et al. (2026), with clinically defined features following FIGO guidelines (Ayres-de-Campos et al., 2015).

Finally, we trained an *InceptionTime* terminal classifier, following McCoy et al. (2024), trained on the final hour of pH-labeled FHR/TOCO tracings at 0.25 Hz with multi-label binary cross-entropy. Because it does not model delivery timing, it provides a pH-risk, not time-to-delivery, baseline. Training details are reported in Appendix D.

### 4.3. External validation on CTU-UHB

We externally validated on the 522 CTU-UHB recordings (Chudáček et al., 2014), resampled to 0.25 Hz and labeled at the same pH thresholds. No CTU data were used for training, selection, or finetuning. Because recordings contain only the final 90 minutes of labor, we evaluated the 0- and 20-minute end-landmarks.

## 5. Results

### 5.1. Cohort Construction and Characteristics

Of 124,777 raw EFM files, 120,403 were retained; 119,261 (99.1%) were preserved in full and 1,142 (0.9%) were truncated to their final 48 hours. After removing 1,180 possible twins, 33,205 patient– lab-time records remained. Delivery-time supervision included 108,046 training and 4,648 validation traces; the pH-labeled cohort included 8,508 training and 2,163 validation traces. The held-out pH-labelled cohort comprised 3,530 patients. Landmark-specific analyses further required an eligible preceding one-hour tracing window, so their denominators vary by landmark. Appendix E illustrates cohort construction.

Most pH values were above 7.20; prevalences below 7.20, 7.15, 7.10, and 7.05 were 20.9%, 9.1%, 3.3%, and 1.3%. Median maternal age was 28.0 years (IQR, 23.0–32.0) and gestational age was 39.3 weeks (IQR, 38.3–40.1). Participants were 62.8% Black, 19.3% White, 7.3% Asian, 4.4% Hispanic or Latino, 0.7% East Indian, and 5.6% other or unknown.

### 5.2. Acidemic Delivery Risk Across Labor

Table 2 reports 1-hour acidemic-delivery macro-average AUROC—the unweighted mean of the separately computed values for each pH threshold—across models and landmarks; the threshold-specific table appears in Appendix F and AP-lift table appears in Appendix G. The marked DeepHit model was best except at one setting, with AUROCs ranging from 0.837 [0.692–0.970] to 0.935 [0.858–0.964].

**Table 1:** Delivery characteristics by cord pH availability. Values are median [IQR].

| Characteristic | pH observed | pH missing |
|---|---|---|
| Patients | 14,737 | 89,110 |
| Maternal age, years | 28.0 [23.0, 32.0] | 29.0 [25.0, 33.0] |
| Gestational age, weeks | 39.3 [38.3, 40.3] | 39.3 [38.3, 40.1] |
| Tracing duration, hours | 13.3 [8.2, 20.0] | 10.9 [4.9, 19.3] |

**Table 2:**
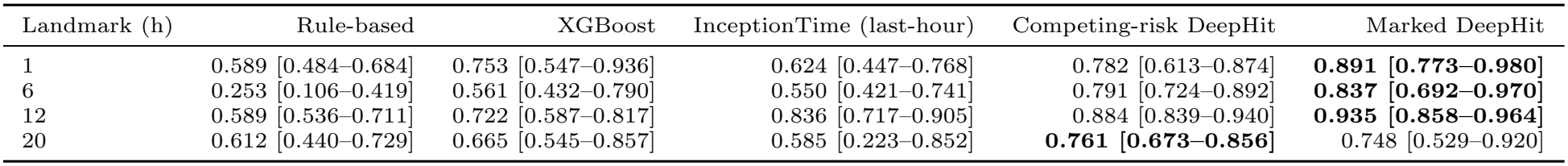
Test-set macro AUROC for acidemic delivery within a 1-hour forecast horizon. The best estimate and interval in each landmark–pH-category row are bolded.

Threshold-specific results showed endpoint-level differences. At the 1-hour landmark, Marked Deep-Hit achieved AUROCs of 0.932, 0.918, and 0.917 for pH *<* 7.10, 7.15, and 7.20, respectively, whereas competing-risk DeepHit led for pH *<* 7.05 (0.843 versus 0.796). At 20 hours, competing-risk Deep-Hit led for pH *<* 7.10, 7.15, and 7.20 (0.755, 0.821, and 0.830), while Marked DeepHit led for pH *<* 7.05 (0.838 versus 0.638). None of the remaining models achieved best performance on the acidemic delivery within one hour task.

Figure 1 shows *pH macro AUROC*, i.e., the average of AUROC across pH thresholds, for acidemic delivery predicted up to 4 hours before delivery. Across methods, average performance declines as the forecast horizon increases. The rule-based method performs below random guessing at the 6-hour labor landmark and near random across the remaining evaluated horizons and landmarks. The InceptionTime terminal model outperformed both rule-based and XGBoost baselines in three settings: 1-, 2-, and 4-hour horizons at the 12-hour landmark. Marked DeepHit performed best in all but one setting, with AUROC above 0.7 across the 1-, 6-, 12-, and 20-hour landmarks and 1-, 2-, and 4-hour horizons.

**Figure 1.**
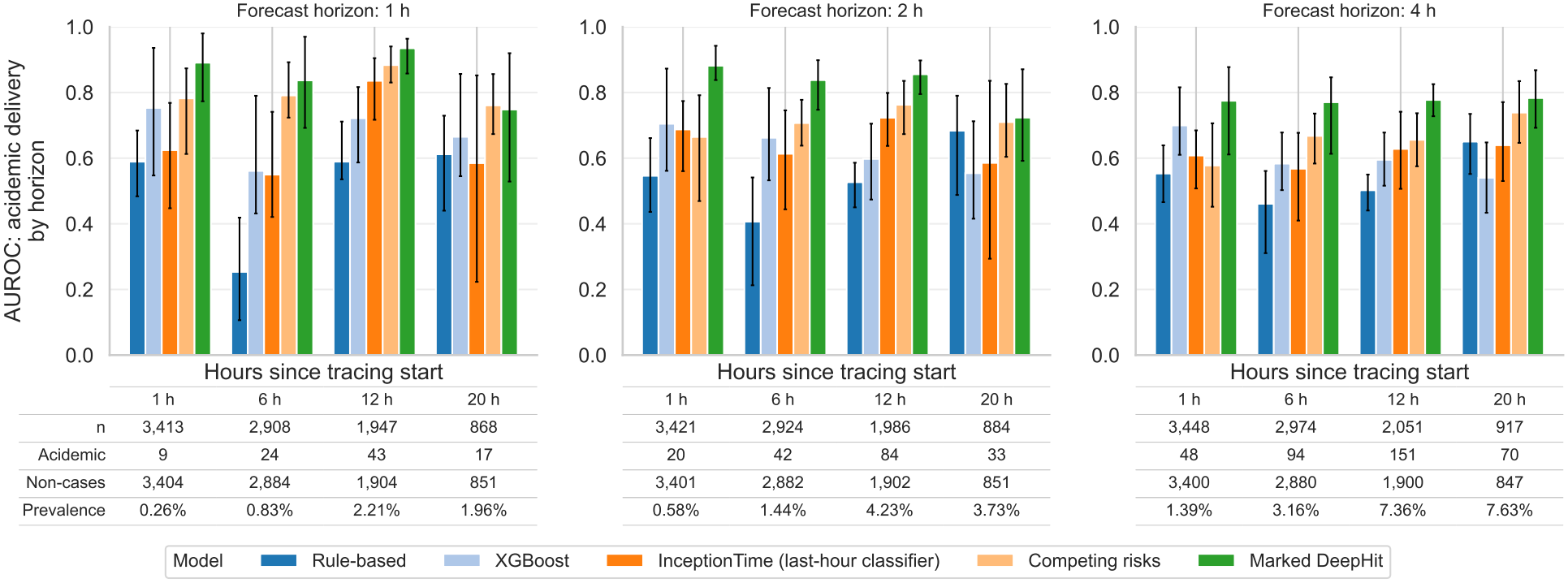
Macro-average AUROC for predicting acidemic delivery within 1-, 2-, and 4-hour horizons, assessed at 1, 6, 12, and 20 hours after monitoring begins. Tables beneath each panel report the number of eligible patients, acidemic cases, non-cases, and acidemia prevalence at different hours since start.

### 5.3. Delivery-Time Discrimination

Figure 2 evaluates acidemia prediction at landmarks before tracing end across all available tracings. AU-ROCs are modest (between 0.6–0.8) and broadly similar across models, with the last-hour InceptionTime classifier performing best at tracing end. Thus, when tracing end is known, models have comparable ability to predict pH thresholds, with performance generally declining farther from the end. Earlier end-landmarks include fewer tracings since short recordings cannot support the required horizon.

**Figure 2.**
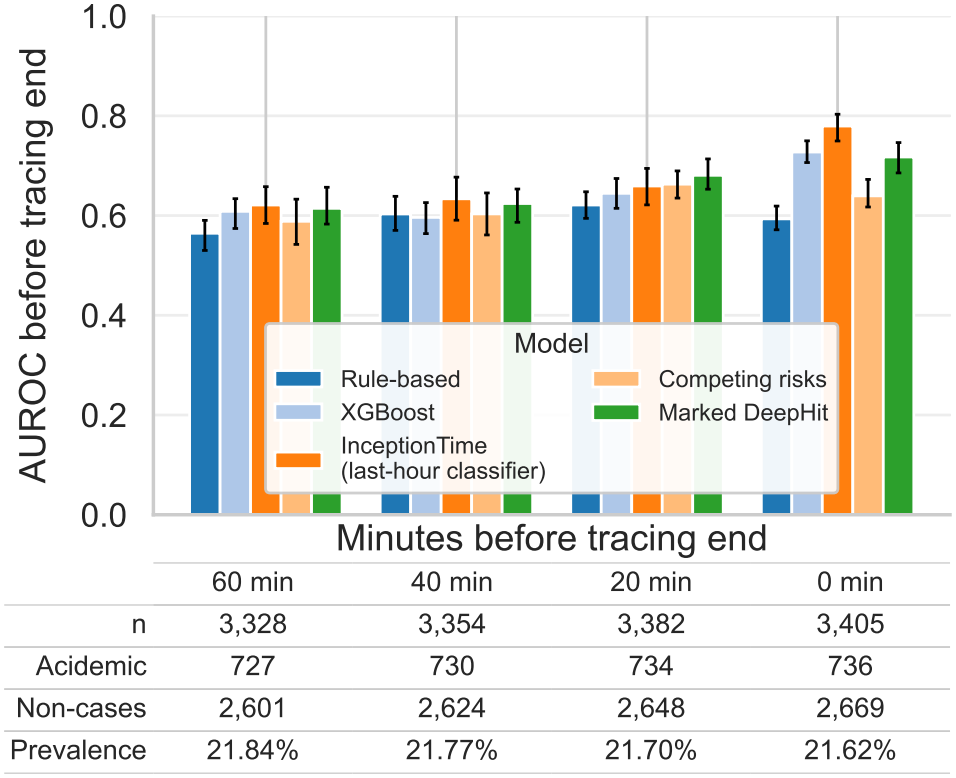
End-anchored evaluation of macro-average AUROC for acidemic delivery across cord-pH thresholds at landmarks before tracing end. The 0-minute landmark corresponds to terminal classification; earlier landmarks assess prediction 20, 40, and 60 minutes before the known tracing end. Counts are unique pH-observed test patients eligible at each landmark with a retrospective window of one hour

Figure 3 reports discrimination for delivery within one hour at elapsed-time landmarks. The Inception-Time terminal model, which does not model time to delivery, performs at chance (AUROC 0.5), whereas both time-to-event models substantially outperform the baselines. Delivery prediction becomes easier at later landmarks, likely because elapsed time provides information about proximity to delivery.

**Figure 3.**
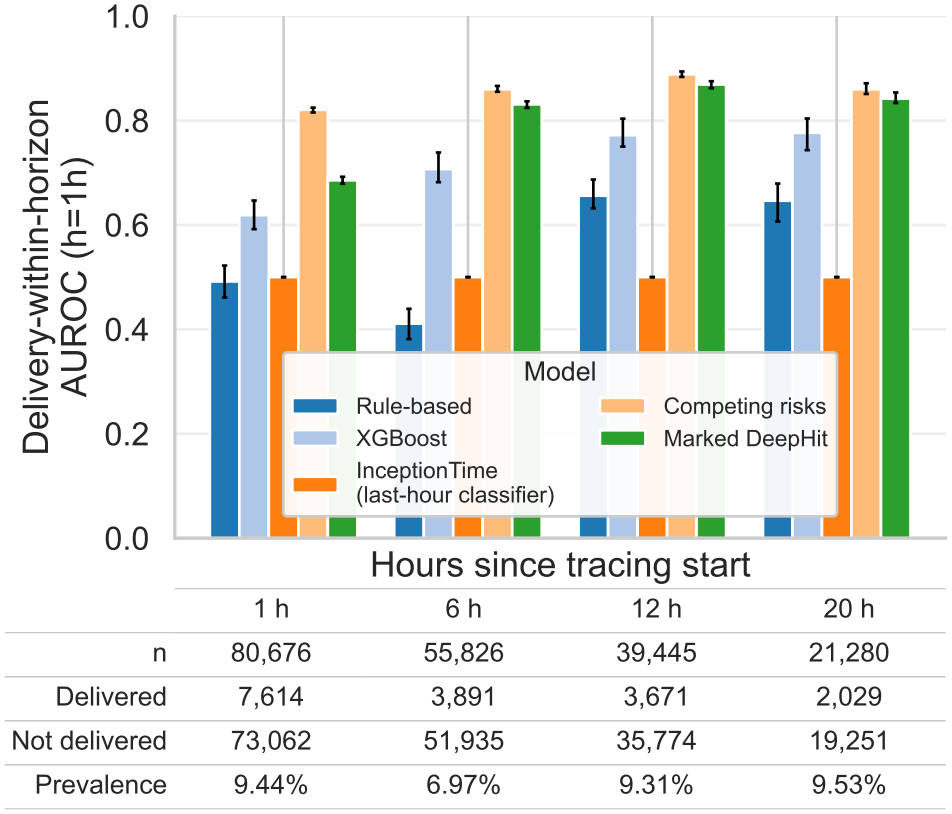
Test-set AUROC for delivery within one hour at elapsed-time landmarks. This is the delivery timing prediction target, irre-spective of cord pH, and therefore includes all eligible test traces rather than only pH-labeled traces. Counts are unique tracings per landmark.

Although acidemia discrimination is modest, the proposed models show strong delivery-timing discrimination—a capability unavailable to terminal classifiers and poorly captured by the baselines. More tracings are available for this outcome since cord-pH measurement is not required. Antolini concordance results are reported in Appendix H.

Marked DeepHit discriminated delivery within one hour with AUROC of 0.685 [0.679–0.692] at the 1-hour landmark, 0.831 [0.824–0.837] at 6 hours, and 0.869 [0.862–0.875] at 12 hours. Its AUROC was 0.842 [0.833–0.854] at the 20-hour landmark, exceeding the corresponding XGBoost AUROC of 0.776 [0.743–0.804].

## 6. Clinical alert operating points

At the 2-hour forecast horizon, validation-selected alert thresholds helps to have a glimpse on clinical alert. We targeted a sensitivity or specificity of *≥*80% and reported test performance on Table 3. For the sensitivity-targeted operating point, test sensitivity ranged from 66.7% to 84.2%, with alert rates of 19.5%–24.9%. PPV increased from 2.4% at the 1-hour landmark to 10.8% at 20 hours, following an increase in event frequency. For the specificity-targeted operating point, test specificity remained close to 80% across landmarks, while sensitivity ranged from 57.6% to 84.2%. Results for all forecast horizons are reported in Table I.1.

**Table 3:** Held-out test performance for Marked DeepHit’s pH *<* 7.2 alerts within a two-hour forecast horizon. Risk thresholds were selected for different operating points using the validation partition and applied unchanged to the test cohort.

| Operating point | Landmark (h) | Sensitivity | Specificity | Events | Alert rate | PPV | Lift |
|---|---|---|---|---|---|---|---|
| Sensitivity ≥ 80% | 1 | 84.2% | 80.9% | 19/3,405 (0.6%) | 19.5% | 2.4% | 4.32 |
|  | 6 | 66.7% | 75.7% | 42/2,886 (1.5%) | 24.9% | 3.9% | 2.67 |
|  | 12 | 71.4% | 80.1% | 84/1,907 (4.4%) | 22.1% | 14.2% | 3.23 |
|  | 20 | 66.7% | 77.8% | 33/853 (3.9%) | 23.9% | 10.8% | 2.79 |
| Specificity ≥ 80% | 1 | 84.2% | 79.8% | 19/3,405 (0.6%) | 20.5% | 2.3% | 4.10 |
|  | 6 | 64.3% | 79.4% | 42/2,886 (1.5%) | 21.3% | 4.4% | 3.02 |
|  | 12 | 72.6% | 79.6% | 84/1,907 (4.4%) | 22.7% | 14.1% | 3.21 |
|  | 20 | 57.6% | 79.0% | 33/853 (3.9%) | 22.4% | 9.9% | 2.57 |

### 6.1. Delivery-risk calibration and operating points

We assessed calibration for Marked DeepHit’s acidemic delivery predictions at the four elapsed-time landmarks and horizons. At the 1-hour horizon, calibrated expected-to-observed ratios were 1.11, 0.79, 0.69, and 1.96 and Brier scores were 0.003, 0.008, 0.020, and 0.019 for 1-, 6-, 12-, and 20-hour landmarks, respectively. Appendix Table J.1 reports all event count, observed prevalence, expected-to-observed ratio, and Brier score at each landmark.

### 6.2. Terminal Discrimination and Delivery-Risk Stratification

The terminal classifier performed best when its end-of-tracing assumption held: at tracing end, it achieves AUROC, macro-averaged across pH thresholds, of 0.779 [0.755–0.810], compared with 0.717 [0.688–0.746] for Marked DeepHit and 0.727 [0.700– 0.751] for XGBoost. Within each elapsed-time landmark, shorter remaining-time bands generally receive higher predicted risks, and the distributions shift lower as more time remains. This descriptive pattern supports that the model captures temporal proximity to delivery. Appendix K address terminal acidemia discrimination.

### 6.3. External Validation

Table 4 reports discrimination for acidemic delivery within a one-hour forecast horizon across elapsed-time landmarks and pH thresholds on the CTU-UHB external dataset. The classification model trained with last hour of tracings achieved the best performance predicting acidemic delivery right at the end of tracings, achieving an AUROC of 0.722 [0.665– 0.767]. When a horizon landmark of 20 minutes is taken into account, the Marked DeepHit achieved the best performance, achieving an AUROC of 0.688 [0.620–0.768].

**Table 4:** External CTU-cohort AUROC at 0 and 20 minutes before delivery.

| Model | Landmark | AUROC |
|---|---|---|
| Rule-based | 0 min | 0.573 [0.518–0.631] |
|  | 20 min | 0.543 [0.482–0.607] |
| XGBoost Cox | 0 min | 0.655 [0.605–0.703] |
|  | 20 min | 0.570 [0.495–0.643] |
| InceptionTime (last hour) | 0 min | **0.722 [0.665-0.767]** |
|  | 20 min | 0.667 [0.592–0.725] |
| Competing-risk DeepHit | 0 min | 0.650 [0.603–0.702] |
|  | 20 min | 0.686 [0.605–0.779] |
| Marked DeepHit | 0 min | 0.702 [0.644–0.743] |
|  | 20 min | **0.688 [0.620-0.768]** |

## 7. Discussion and Conclusion

This work reframes intrapartum AI-EFM for prospective deployment and demonstrates the importance of landmark-time evaluation. Classifiers trained only on the final-hour tracings underperform elsewhere during labor, suggesting real-world analyses must account for time to delivery. Deep time-to-event models like Marked DeepHit accurately rank near-term delivery risk across landmarks and exhibit competitive acidemia discrimination near tracing end.

Our results also distinguish delivery-time discrimination from acidemia discrimination. Delivery within a one-hour horizon was predicted substantially better than acidemia at earlier landmarks, demonstrating that delivery timing is an informative and accessible target. During ongoing labor, neither remaining time to delivery nor eventual cord pH is known. The operational forecast is therefore whether an acidemic delivery is likely to occur in the next one, two, or four hours, using the preceding hour of CTG and elapsed monitoring time. This risk may support surveillance or escalation, but is not intended to replace clinical judgment or mandate intervention.

Performance was not uniform across landmarks. At 20 hours, fewer pH-labeled deliveries, especially at severe-acidemia thresholds, yield wider intervals; longer horizons may also be more sensitive to intervening clinical events and physiologic change. By contrast, the terminal InceptionTime classifier performs best at tracing end because it is trained specifically on final-hour windows, an assumption available only retrospectively and therefore an optimistic terminal-use comparison. The CTU-UHB recordings similarly contain only approximately 60–90 minutes near delivery and lack elapsed labor time, so they support terminal discrimination but cannot assess the earlier landmark predictions motivating this work.

Published terminal EFM studies provide useful context, but address a different decision setting. Our models learn from CTG signals and are trained and evaluated throughout labor. More broadly, this is a dynamic landmark-prediction problem from longitudinal clinical signals, akin to repeatedly updating deterioration risk from evolving ICU or inpatient monitoring data.

Several limitations of our study should be noted. External validation was limited to end-of-tracing recordings available publicly; future work should externally validate elapsed-time landmark performance. This retrospective analysis uses only FHR and TOCO and does not establish clinical benefit or account for clinical information available to care teams. Cord pH was missing for some deliveries; although the site has a uniform collection policy and our loss retains delivery-time supervision, missingness may still be related to unmeasured confounders like fetal status or clinical concern. The pH timing rule requires the laboratory order to occur after and within 30 minutes of the final EFM timestamp. It improves temporal linkage but does not establish that monitoring stopped exactly at delivery; the corresponding timing check was unavailable for CTU-UHB. Although arterial cord pH is clinically meaningful and associated with adverse short- and long-term outcomes, it is an imperfect surrogate for those outcomes; and we do not establish that predicting acidemia improves neonatal outcomes. Prospective multicenter studies should assess alert burden, clinical decisions, and additional outcomes.

Overall, time-to-event EFM models may offer a more deployable and clinically meaningful target than end-of-tracing classification alone by providing a prospective delivery-risk signal while updating acidemia risk as delivery approaches.

## Data Availability

We use data from the CTU-UHB repository and a multi-site academic health system (University of Pennsylvania Health System). Access to CTU-UHB is openly available under the terms of the data use agreement on Physionet. Code for this paper is available in the following repository: https://github.com/cavalab/deep-tte-intrapartum-fetal-monitoring

https://physionet.org/physiobank/database/ctu-uhb-ctgdb/

## Acknowledgments

This work was partially supported by the Eunice Kennedy Shriver National Institute Of Child Health & Human Development of the National Institutes of Health under Award Number R01HD119210. The content is solely the responsibility of the authors and does not necessarily represent the official views of the National Institutes of Health. The authors acknowledge the use of AI for code development, math checking and copy-editing.

## Appendix A. Extended Methods

This appendix specifies the loss terms omitted from the main Methods section.

For the discrete-time heads, 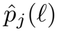 is the probability mass assigned to delivery-time bin *l* and cause *j*. It is obtained from a softmax over cause–bin cells, augmented by one extra “no event within the grid” cell:

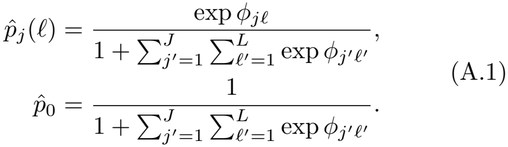

Here, 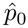is the probability of no delivery within the discretized grid.

### A.1. Competing risks: partial-label loss

For a batch of *N* landmark examples, let *l*_*i*_ = *l*(*T*_*i*_). Let *e*_*i*_ = *E*_*i*_ when *R*_*i*_ = 1, and reserve *e*_*i*_ = 0 for a delivery whose pH was not measured. Split the batch into *O*= {*i* : *e*_*i*_*≠*0} and ℳ= {*i* : *e*_*i*_ = 0 }. We set a loss component associated with an empty subset to zero. For pH-observed rows, the cause-specific negative log-likelihood is:

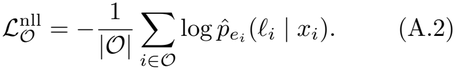

Let 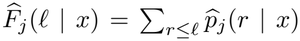 denote the cause-specific cumulative incidence. Because delivery is observed for every retained pregnancy, the comparability mask selects pairs in which *i* delivers strictly before *i*^*′*^, namely 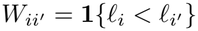. The cause-specific ranking term is:

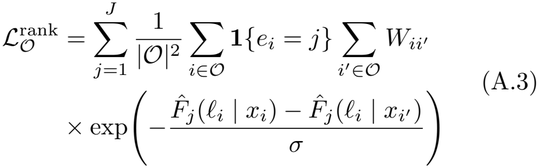

This smooth surrogate penalizes an earlier delivery of cause *j* when it receives lower cumulative incidence at its delivery time than a later delivery. Here, *σ >* 0 is a ranking-temperature hyperparameter; smaller values impose a steeper penalty for incorrectly ordered pairs.

For pH-missing rows, the delivery bin is known but the cause is not. Their loss marginalizes the joint mass over causes and ranks delivery times using the all-cause cumulative incidence. Let 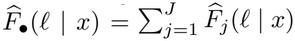 denote that cumulative incidence.

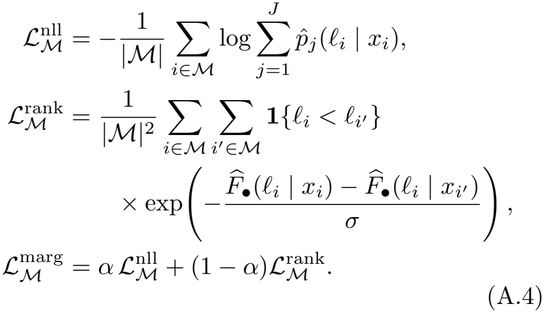

The partial-label objective in Eq. (A.4) lets pH-missing samples train delivery timing without contributing pH-category supervision.

The complete competing-risks objective weights the two populations by their shares of the batch:

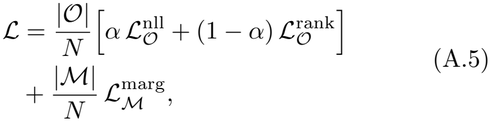

Here, *α ∈* [0, 1] trades off the likelihood and ranking terms for both pH-observed and pH-missing rows.

### A.2 Marked point process: observed-mark loss

The marked model applies the ordinary single-risk DeepHit delivery loss to all rows, as described in the main text. Only rows with observed pH contribute a mark target. Writing 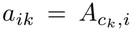 for the cumulative threshold label, the additional mark loss is a binary cross-entropy over thresholds evaluated at the observed delivery bin:

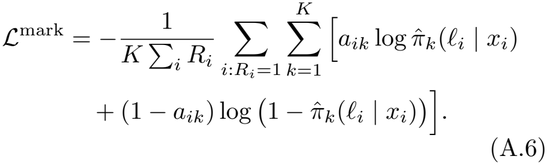

The complete marked objective is given in the main text by the delivery loss plus *w*_mark_*L*mark, with *w*_mark_ = 1 by default. Thus, the mask keeps pH-missing deliveries in the timing loss and out of the pH loss without a separate marginalization term.

## Appendix B. Deep time to event network configuration

The encoder input consists of the `toco` and `fecg` channels, an elapsed-time channel, and a imputation observation channel. The input window contains 900 samples, corresponding to 3, 600 seconds (1 hour) at a sampling frequency of 0.25 Hz.

Missing values are forward-filled after chunk selection. Chunks with a raw missingness fraction above 0.30 are considered ineligible, while the observation-mask channels preserve information about pre-imputation observability. The model is optimized using Adafactor with *β*_2_ decay = *−*0.8, *ϵ* = (None, 10^*−*3^), a relative-step scale/cap of *d* = 1.0, an update clipping threshold of 1.0, and no weight decay. The learning rate follows a repeated cosine annealing schedule (Loshchilov and Hutter, 2017) with an initial value of 1 *×*10^*−*2^, a linear warm-up over 5% of the planned optimizer updates, and a decay to 1% of the initial value (1 *×* 10^*−*2^).

Training is performed for a maximum of 100 epochs with an early-stopping patience of 30 epochs. The model weights corresponding to the lowest validation loss are always restored before evaluation. The batch size is 2048 chunks/PID examples.

The DeepHit loss uses *α* = 0.5 and *σ* = 0.2 throughout. The marked loss uses a multiplier of 1.0 and masked binary cross-entropy at the observed delivery-time bin. The competing-risks loss uses the partial-label competing DeepHit formulation with a missing-delivery loss multiplier of 1.0.

The cause-specific competing-risks head consists of one independent head per cause, with hidden-layer widths of (128, 64), batch normalization enabled, and dropout of 0.2.

## Appendix C. Parameter sweep

Ismail Fawaz et al. (2020) adapt the receptive field of a network for time-series data as its field of view for detecting patterns. For a sequential convolutional path with unit strides and no dilation, its receptive field is 1 + ∑*i*(*k*_*i*_ *−*1). In our InceptionTime architecture, each residual block contains three sequential modules, and the longest path through each module uses the largest kernel *k*_max_ from its parallel convolutional branches. Thus, for *d*_blocks_ residual blocks, the receptive field is

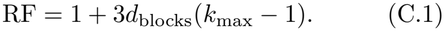

that increasing filter length grows the RF linearly in *k*, while increasing depth grows it only linearly in *d* with a much smaller slope — and that on real UCR datasets, deeper networks tend to overfit without improving accuracy, whereas longer filters reliably help up to the point where they start to overfit smaller datasets.

Because our series are moderately long (900 samples corresponding to 1 hour of tracing data collected at 0.25 sample rate, 2 channels), we focus the optimization to two hyperparameters directly tied to the RF: the Inception filter-length triple, scaled from the paper’s default {10, 20, 40}, and corrected to odd values as required by the implementation’s padding scheme (yielding {9, 19, 39}, {5, 9, 19}, and {19, 39, 79} ), and the network depth, expressed as the number of residual blocks of three Inception modules each (*d*_blocks_ *∈* {1, 2, 3}, spanning the paper’s default of 2 blocks / 6 modules).

InceptionTime consists of a set of modules that apply a channel bottleneck, parallel convolutions at several kernel widths, and a parallel max-pooling branch; modules are stacked with residual connections and terminated by global average pooling. We configured the encoder to produce a 128-dimensional embedding.

In our implementation of InceptionTime, each depth corresponds to the number of InceptionTime blocks, where each block consists of three Inception modules, each containing three parallel convolutions with different kernel sizes and one max-pooling layer, followed by a residual shortcut. We set *n*_filters_ = 32, *n*_bottleneck_ = 32 at InceptionTime’s published defaults.

**Table C.1:** Receptive field (RF; calculated by Eq. C.1) and fraction of the 900-sample series it covers, for each swept combination of filter-length triple *{k*_1_, *k*_2_, *k*_3_*}* and depth *d*_blocks_ (each block = 3 Inception modules). Bold marks the InceptionTime default configuration.

|  | Depth (blocks / modules) |  |  |
|---|---|---|---|
| Kernel sizes | 1 / 3 | **2 / 6** | 3 / 9 |
| {5, 9, 19} | 55 (6%) | 109 (12%) | 163 (18%) |
| **{9, 19, 39}** | 115 (13%) | **229 (25%)** | 343 (38%) |
| {19, 39, 79} | 235 (26%) | 469 (52%) | 703 (78%) |

### C.1 Train sampling regime

The training-sampling regime was treated as an experimental factor because it changes the distribution of observable landmarks presented during training and can therefore affect joint delivery-time and pH-mark prediction, especially given that unlabeled data is using mixed with labeled data. Training traces are streamed lazily, with one or more selected chunks per patient per epoch according to a sampling regime.

We considered the following policies. The `stratified_elapsed` regime divided each patient’s observable tracing interval into four elapsed-time strata and rotated patients through these strata across epochs, balancing time since tracing start without using delivery information. The `terminal_balanced` regime alternated the four elapsed-time strata with four delivery-relative strata covering 0–10, 10–30, 30–50, and 50–70 minutes before delivery. This provided a balanced mixture of ordinary elapsed-time examples and examples near the fixed 0-, 20-, 40-, and 60-minute end landmarks. The `quartiles_plus_terminal` regime sampled five chunks per patient per epoch: one eligible chunk from each of four elapsed-time quartiles and one additional eligible chunk ending within 20 minutes of delivery. Finally, the `all_strata` regime enumerated every eligible, aligned, non-overlapping chunk available for a patient, and used 4 chunks sampled uniformly for each patient. For the delivery-relative policies, delivery time was used only to select retrospective training chunks and was never supplied as a model input.

### C.2 Model sizes

Table C.2 reports the number of trainable parameters for each configuration evaluated in the architecture sweep. The configurations are grouped by prediction mode, network depth, and Inception module kernel-size triple. The parameter count increases with both network depth and kernel size, while the competing-risk configuration has a larger number of parameters than the corresponding classifier configuration due to its cause-specific prediction heads.

### C.3 Model selection

For each neural model family, we selected a single configuration using the validation set only. Candidate configurations varied in InceptionTime architecture and training-sampling regime. We ranked candidates by mean AUROC, macro-averaged across pH thresholds and all available acidemic-delivery forecast-horizon and end-landmark evaluations, and retained the highest-scoring configuration within each family for held-out test evaluation. For Marked DeepHit, this procedure selected a forward-filled InceptionTime encoder with three inception blocks and kernel sizes {9, 19, 39} (receptive field 343 samples; 38% of the input window), trained with the `terminal_balanced` regime. For Competing-risk DeepHit, this procedure selected a forward-filled InceptionTime encoder with three inception blocks and kernel sizes {9, 19, 39}, trained with the `quartiles _plus_terminal` regime. The rule-based, XGBoost, and last-hour classifier baselines were fixed comparators and were not selected from this sweep.

**Table C.2:**
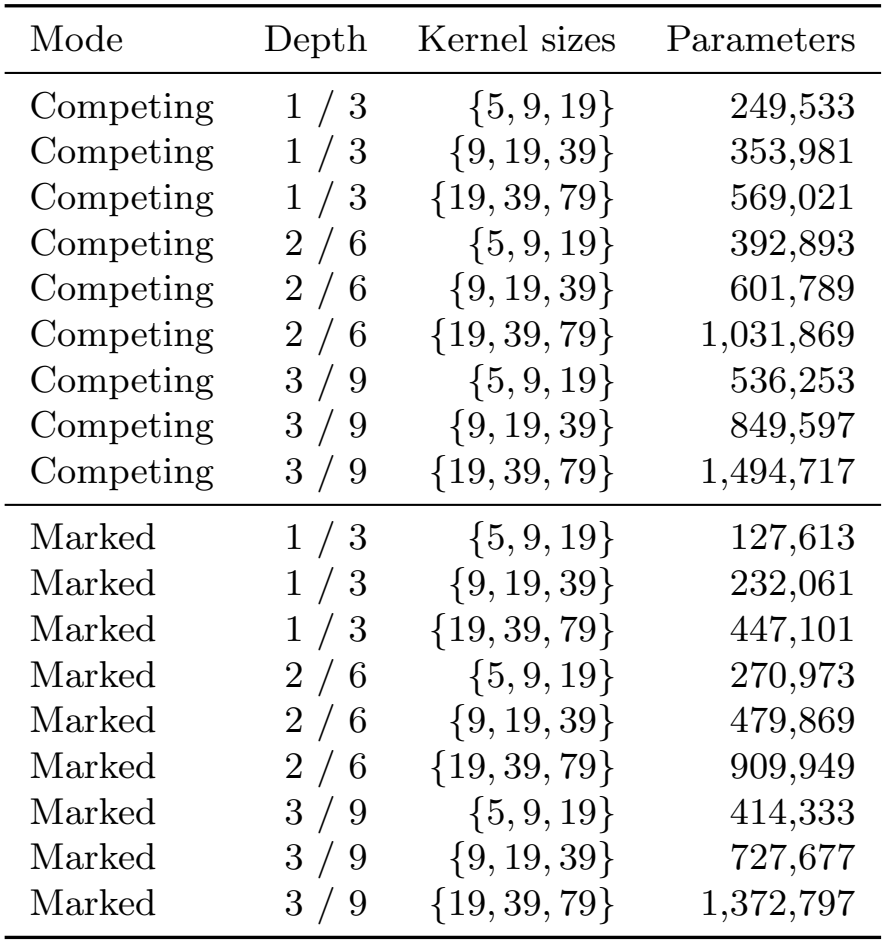
Number of trainable parameters for each configuration in the architecture sweep. Configurations are grouped by prediction mode, depth, and Inception module kernel-size triple.

## Appendix D. Last-hour model

The last-hour model used a two-channel Inception-Time classifier operating on 60-minute windows of fetal heart rate and uterine activity sampled at 0.25 Hz. It was trained using the same patient cohort, labels, and train–validation–test splits as the time-to-event (TTE) models, but using only the final hour of each tracing.

The network consisted of two Inception blocks, each containing three sequential Inception modules with residual connections. Each convolutional branch used 32 filters, with 32-channel bottleneck convolutions and kernel sizes of (39, 19, 9) samples, together with a three-wide max-pooling branch. Global average pooling produced a 128-dimensional representation followed by a linear output layer.

The model produced four logits corresponding to four cord-pH thresholds. It was trained as a multi-label classifier using binary cross-entropy with logits. Inputs were scaled and missing values were forward-filled; windows with more than 30% missing physiological values were excluded. The label configuration used a 30-minute laboratory-order delay and horizon zero. Because the model was intended for prediction from the final hour of the tracing, elapsed time since tracing start was not provided as an input channel.

Training used the Adam optimizer with an initial learning rate of 10^*−*4^ and a minimum learning rate of 10^*−*7^, a batch size of 64, and a maximum of 200 epochs. Early stopping was applied when validation AUROC did not improve for 30 epochs.

## Appendix E. Data Processing

Figure E.1 gives a breakdown of the preprocessing and filtering steps leading to the delivery time and pH labeled cohorts used for training and evaluation.

## Appendix F. Threshold-specific AUROCs

Table F.1 report one-hour acidemic-delivery AUROC across models, landmarks, and pH thresholds. Marked DeepHit had the highest AUROC point estimate in 12 of 20 landmark–pH-category rows, ranging from 0.673 [0.469–0.877] to 0.997 [0.993–0.999]. At the 1-hour landmark, it achieved AUROC, macroaveraged across pH thresholds, of 0.891 [0.773–0.980] and pH *<* 7.1 AUROC of 0.932 [0.886–0.996]. At 12 hours, its AUROC, macro-averaged across pH thresholds, was 0.935 [0.858–0.964], and its pH *<* 7.05 AUROC was 0.997 [0.993–0.999]. The remaining eight rows favored competing-risk DeepHit; at 20 hours, it exceeded Marked DeepHit for macro-average across pH thresholds (0.825 vs. 0.748) and pH thresholds of 7.1 (0.825 vs. 0.688), 7.15 (0.826 vs. 0.673), and 7.2 (0.849 vs. 0.794).

**Figure E.1:**
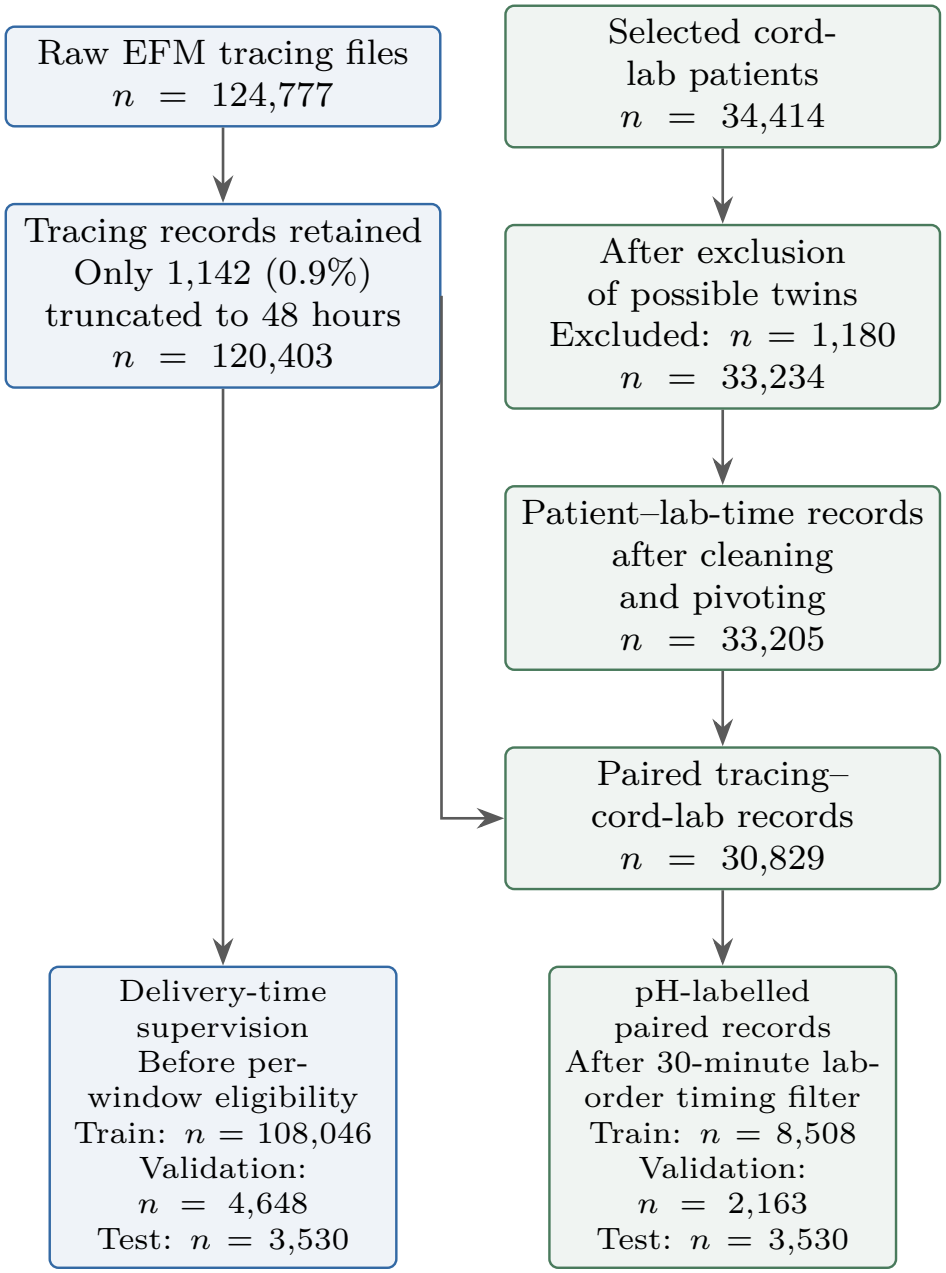
STROBE-style flow diagram of cohort construction from the completed tracing and laboratory preprocessing logs. The pH-labeled branch applies the 30-minute laboratory-order timing criterion, whereas the delivery-time branch retains pH-missing traces for timing supervision.

## Appendix G. AP-lift metrics

Table G.1 reports Average Precision lift (AP lift, prevalence normalized average precision) for acidemic delivery across model families, elapsed landmark times, and pH thresholds, using the same 1-hour forecast horizon as the primary AUROC analysis. AP lift is average precision relative to the observed out-come prevalence, such that a value of 1 indicates prevalence-level performance and larger values indicate greater enrichment of acidemic deliveries among high-risk predictions. The AP lift measures the amount of times the model detects case in relation to the base prevalence.

**Table F.1:** Test-set AUROC for acidemic delivery within a 1-hour forecast horizon. The best estimate and interval in each landmark–pH-category row are bolded.

| Landmark (h) | pH category | Rule-based | XGBoost | InceptionTime (last-hour) | Competing-risk DeepHit | Marked DeepHit |
|---|---|---|---|---|---|---|
| 1 | Macro-average | 0.589 [0.484–0.684] | 0.753 [0.547–0.936] | 0.624 [0.447–0.768] | 0.782 [0.613–0.874] | **0.891 [0.773–0.980]** |
|  | pH < 7.05 | 0.624 [0.454–0.797] | 0.655 [0.353–0.950] | 0.464 [0.243–0.686] | **0.843 [0.737–0.947]** | 0.796 [0.729–0.870] |
|  | pH < 7.1 | 0.706 [0.557–0.797] | 0.745 [0.510–0.985] | 0.621 [0.398–0.747] | 0.739 [0.641–0.840] | **0.932 [0.886–0.996]** |
|  | pH < 7.15 | 0.654 [0.458–0.859] | 0.799 [0.607–0.985] | 0.756 [0.611–0.912] | 0.782 [0.641–0.904] | **0.918 [0.771–0.996]** |
|  | pH < 7.2 | 0.373 [0.167–0.566] | 0.814 [0.650–0.970] | 0.656 [0.401–0.875] | 0.765 [0.661–0.890] | **0.917 [0.809–0.991]** |
| 6 | Macro-average | 0.253 [0.106–0.419] | 0.561 [0.432–0.790] | 0.550 [0.421–0.741] | 0.791 [0.724–0.892] | **0.837 [0.692–0.970]** |
|  | pH < 7.05 | 0.275 [0.027–0.526] | 0.095 [0.029–0.168] | 0.444 [0.424–0.461] | 0.682 [0.663–0.698] | **0.850 [0.837–0.861]** |
|  | pH < 7.1 | 0.263 [0.062–0.455] | 0.604 [0.353–0.823] | 0.677 [0.392–0.975] | 0.805 [0.669–0.884] | **0.824 [0.591–0.997]** |
|  | pH < 7.15 | 0.252 [0.134–0.386] | 0.757 [0.635–0.855] | 0.542 [0.342–0.770] | 0.808 [0.647–0.897] | **0.817 [0.595–0.944]** |
|  | pH < 7.2 | 0.224 [0.140–0.331] | 0.789 [0.710–0.869] | 0.537 [0.378–0.662] | **0.869 [0.739–0.959]** | 0.857 [0.735–0.957] |
| 12 | Macro-average | 0.589 [0.536–0.711] | 0.722 [0.587–0.817] | 0.836 [0.717–0.905] | 0.884 [0.830–0.940] | **0.935 [0.858–0.964]** |
|  | pH < 7.05 | 0.157 [0.034–0.227] | 0.627 [0.601–0.654] | 0.972 [0.953–0.989] | 0.860 [0.734–0.980] | **0.997 [0.993–0.999]** |
|  | pH < 7.1 | 0.784 [0.654–0.885] | 0.673 [0.447–0.891] | 0.808 [0.617–0.971] | 0.917 [0.851–0.972] | **0.954 [0.877–0.995]** |
|  | pH < 7.15 | 0.708 [0.564–0.807] | 0.816 [0.688–0.921] | 0.823 [0.725–0.912] | 0.887 [0.786–0.965] | **0.908 [0.813–0.980]** |
|  | pH < 7.2 | 0.709 [0.642–0.773] | 0.771 [0.696–0.837] | 0.739 [0.663–0.813] | 0.870 [0.800–0.903] | **0.881 [0.823–0.921]** |
| 20 | Macro-average | 0.612 [0.440–0.729] | 0.665 [0.545–0.857] | 0.585 [0.223–0.852] | **0.761 [0.673–0.856]** | 0.748 [0.529–0.920] |
|  | pH < 7.05 | 0.500 [0.500–0.500] | 0.500 [0.500–0.500] | 0.464 [0.000–0.938] | 0.638 [0.443–0.838] | **0.838 [0.677–0.997]** |
|  | pH < 7.1 | 0.735 [0.420–0.975] | 0.741 [0.553–1.000] | 0.618 [0.003–0.944] | **0.755 [0.667–0.889]** | 0.688 [0.312–0.993] |
|  | pH < 7.15 | 0.591 [0.300–0.752] | 0.738 [0.465–0.970] | 0.697 [0.411–0.953] | **0.821 [0.741–0.910]** | 0.673 [0.469–0.877] |
|  | pH < 7.2 | 0.621 [0.513–0.755] | 0.682 [0.543–0.781] | 0.561 [0.384–0.698] | **0.830 [0.750–0.893]** | 0.794 [0.691–0.868] |

## Appendix H. Antolini Concordance Index

We report Antolini concordance (Antolini et al., 2005) as a complementary event-time analysis. For the separate delivery-timing prediction task, Figure H.1 summarizes Antolini concordance, macro-averaged across pH thresholds, at elapsed-time landmarks.

For Antolini concordance, macro-averaged across pH thresholds, Marked DeepHit increased from 0.573 [0.526–0.619] at 1 hour to 0.796 [0.756–0.831] at 20 hours.

## Appendix I. Clinical pH *<* 7.2 alert operating points

Table I.1 reports the remaining validation-selected operating points for one- and four-hour forecast horizons.

**Figure H.1:**
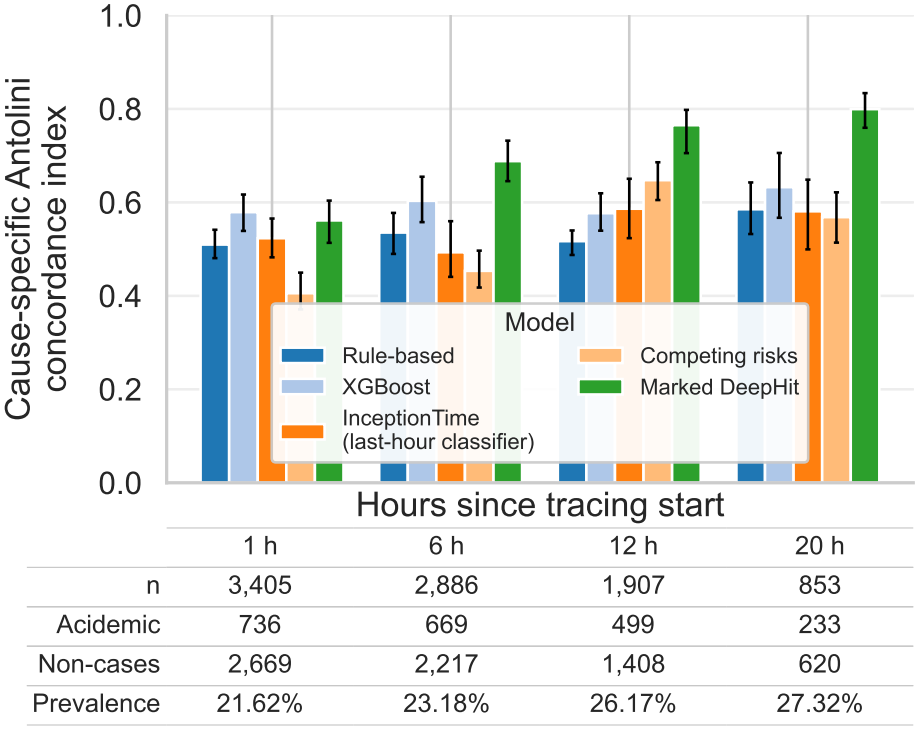
Antolini concordance, macro-averaged across pH thresholds, at elapsed-time landmarks on the test set.

## Appendix J. Delivery-risk calibration

Table J.1 summarizes raw and validation-recalibrated delivery-within-one-hour risk on the held-out pH-observed test cohort.

“‘latex “‘

**Table G.1:** Test-set AP lift for acidemic delivery within a 1-hour forecast horizon. AP lift is average precision divided by outcome prevalence; a value of 1 corresponds to prevalence-level performance. Horizons of 2 and 4 hours are omitted. The best estimate and 95% confidence interval in each landmark–pH-category row are bolded.

| Landmark (h) | pH category | Rule-based | XGBoost | InceptionTime (last-hour) | Competing-risk DeepHit | Marked DeepHit |
|---|---|---|---|---|---|---|
| 1 | Macro-average | 1.5 [1.2–3.0] | **33.2 [4.1–395.9]** | 2.4 [1.5–11.8] | 4.0 [2.5–9.3] | 24.9 [11.1–112.2] |
|  | pH < 7.05 | 1.4 [1.2–2.9] | 5.2 [1.5–20.4] | 1.4 [1.2–3.2] | **6.1 [3.8–17.7]** | 3.6 [3.2–7.5] |
|  | pH < 7.1 | 1.9 [1.3–2.9] | 10.3 [1.6–124.1] | 1.6 [1.4–4.5] | 3.7 [2.0–19.0] | **18.2 [6.7–88.5]** |
|  | pH < 7.15 | 1.7 [0.9–5.1] | **101.4 [2.4–391.4]** | 4.0 [1.6–20.7] | 3.4 [2.0–10.4] | 48.3 [18.6–203.4] |
|  | pH < 7.2 | 0.9 [0.8–1.6] | 15.8 [5.3–41.0] | 2.7 [1.4–9.0] | 2.9 [1.8–7.4] | **29.7 [13.3–118.2]** |
| 6 | Macro-average | 0.8 [0.8–1.0] | 3.6 [2.3–10.0] | 6.4 [1.4–86.8] | 6.0 [4.2–16.3] | **21.9 [8.5–138.0]** |
|  | pH < 7.05 | 0.9 [0.8–1.5] | 0.8 [0.8–1.2] | 1.8 [1.7–1.9] | 3.1 [2.9–3.4] | **6.7 [6.2–7.2]** |
|  | pH < 7.1 | 0.7 [0.7–1.1] | 1.6 [1.0–3.9] | 17.2 [1.3–131.5] | 3.1 [2.6–6.2] | **44.8 [3.4–263.2]** |
|  | pH < 7.15 | 0.7 [0.7–0.9] | 4.2 [2.4–9.5] | 3.8 [0.8–17.9] | 3.7 [2.4–7.8] | **14.7 [6.6–41.7]** |
|  | pH < 7.2 | 0.7 [0.7–0.8] | 7.7 [3.4–19.7] | 2.6 [1.1–10.6] | 13.9 [6.6–35.5] | **21.4 [7.4–41.8]** |
| 12 | Macro-average | 3.9 [1.5–10.7] | 7.9 [3.1–22.0] | 12.2 [4.1–32.7] | 14.9 [7.6–30.1] | **70.1 [19.1–167.7]** |
|  | pH < 7.05 | 0.8 [0.8–1.1] | 1.8 [1.7–2.8] | 28.5 [18.9–76.5] | 12.0 [3.7–45.9] | **169.5 [94.7–508.6]** |
|  | pH < 7.1 | 2.6 [1.6–6.1] | 5.8 [1.4–56.9] | 7.1 [2.4–23.2] | 13.6 [5.2–54.6] | **72.7 [19.3–347.5]** |
|  | pH < 7.15 | 9.2 [1.4–35.2] | 18.8 [5.9–44.3] | 8.7 [4.3–18.6] | 21.2 [7.2–62.5] | **27.0 [10.4–68.3]** |
|  | pH < 7.2 | 3.0 [1.6–6.9] | 5.1 [3.0–9.5] | 4.7 [2.7–8.8] | **12.9 [8.1–19.7]** | 11.2 [7.2–17.4] |
| 20 | Macro-average | 1.7 [1.1–5.3] | 14.7 [1.8–109.6] | 3.9 [1.3–11.0] | 3.1 [2.6–7.5] | **15.0 [2.1–87.9]** |
|  | pH < 7.05 | 1.0 [1.0–1.0] | 1.0 [1.0–1.0] | 3.8 [1.0–15.0] | 2.3 [1.7–6.1] | **39.7 [2.9–246.4]** |
|  | pH < 7.1 | 2.9 [1.2–19.3] | **48.8 [1.8–426.5]** | 4.6 [1.0–20.1] | 2.7 [2.3–7.6] | 12.0 [1.4–81.8] |
|  | pH < 7.15 | 1.5 [0.8–6.1] | **6.8 [1.7–28.2]** | 5.2 [1.9–18.1] | 3.2 [2.5–7.5] | 3.8 [1.3–17.6] |
|  | pH < 7.2 | 1.6 [1.0–3.6] | 2.3 [1.5–4.4] | 1.9 [1.1–4.7] | 4.2 [2.5–10.3] | **4.4 [2.2–10.6]** |

## Appendix K. Terminal discrimination curves

These terminal analyses assume that tracing end is known and are therefore retrospective comparisons rather than prospective landmark predictions.

### K.1. Delivery-risk distributions

Appendix Figure K.3 displays validation-only calibrated delivery-within-one-hour risks by observed remaining-time band on a shared 0–1 scale. The risk distributions below complement the calibration curves by showing how validation-only recalibrated delivery risk varies with observed remaining time.

**Table I.1:** Supplementary held-out test performance for Marked DeepHit pH *<* 7.2 alerts at one-, two-, and four-hour forecast horizons. Risk thresholds were selected using validation data and applied unchanged to the test cohort. Event frequency is reported as test events/test patients (percentage).

| Operating point | Horizon (h) | Landmark (h) | Sensitivity | Specificity | Events | Alert rate | PPV | Lift |
|---|---|---|---|---|---|---|---|---|
| Validation sensitivity ≥ 80% | 1 | 1 | 77.8% | 89.8% | 9/3,405 (0.3%) | 10.3% | 2.0% | 7.52 |
|  |  | 6 | 83.3% | 67.7% | 24/2,886 (0.8%) | 32.7% | 2.1% | 2.55 |
|  |  | 12 | 74.4% | 83.9% | 43/1,907 (2.3%) | 17.5% | 9.6% | 4.26 |
|  |  | 20 | 70.6% | 73.9% | 17/853 (2.0%) | 27.0% | 5.2% | 2.62 |
|  | 2 | 1 | 84.2% | 80.9% | 19/3,405 (0.6%) | 19.5% | 2.4% | 4.32 |
|  |  | 6 | 66.7% | 75.7% | 42/2,886 (1.5%) | 24.9% | 3.9% | 2.67 |
|  |  | 12 | 71.4% | 80.1% | 84/1,907 (4.4%) | 22.1% | 14.2% | 3.23 |
|  |  | 20 | 66.7% | 77.8% | 33/853 (3.9%) | 23.9% | 10.8% | 2.79 |
|  | 4 | 1 | 76.6% | 60.3% | 47/3,405 (1.4%) | 40.2% | 2.6% | 1.91 |
|  |  | 6 | 77.7% | 46.7% | 94/2,886 (3.3%) | 54.1% | 4.7% | 1.43 |
|  |  | 12 | 78.1% | 63.8% | 151/1,907 (7.9%) | 39.5% | 15.7% | 1.98 |
|  |  | 20 | 81.4% | 61.4% | 70/853 (8.2%) | 42.1% | 15.9% | 1.93 |
| Validation specificity ≥ 80% | 1 | 1 | 77.8% | 80.7% | 9/3,405 (0.3%) | 19.4% | 1.1% | 4.01 |
|  |  | 6 | 75.0% | 79.7% | 24/2,886 (0.8%) | 20.7% | 3.0% | 3.62 |
|  |  | 12 | 76.7% | 78.7% | 43/1,907 (2.3%) | 22.5% | 7.7% | 3.40 |
|  |  | 20 | 58.8% | 80.1% | 17/853 (2.0%) | 20.6% | 5.7% | 2.85 |
|  | 2 | 1 | 84.2% | 79.8% | 19/3,405 (0.6%) | 20.5% | 2.3% | 4.10 |
|  |  | 6 | 64.3% | 79.4% | 42/2,886 (1.5%) | 21.3% | 4.4% | 3.02 |
|  |  | 12 | 72.6% | 79.6% | 84/1,907 (4.4%) | 22.7% | 14.1% | 3.21 |
|  |  | 20 | 57.6% | 79.0% | 33/853 (3.9%) | 22.4% | 9.9% | 2.57 |
|  | 4 | 1 | 63.8% | 80.2% | 47/3,405 (1.4%) | 20.4% | 4.3% | 3.13 |
|  |  | 6 | 46.8% | 79.9% | 94/2,886 (3.3%) | 21.0% | 7.3% | 2.23 |
|  |  | 12 | 55.6% | 80.1% | 151/1,907 (7.9%) | 22.8% | 19.4% | 2.44 |
|  |  | 20 | 50.0% | 79.7% | 70/853 (8.2%) | 22.7% | 18.0% | 2.20 |

**Figure K.1:**
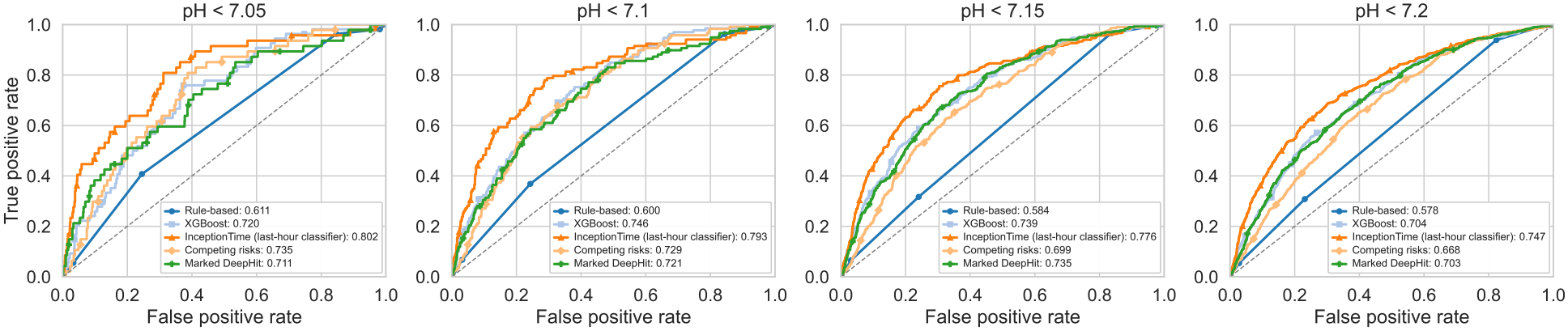
Test-set ROC curves for acidemia at tracing end. Panels are labeled by the cord-pH endpoint.

**Table J.1:** Held-out calibration of Marked DeepHit’s pH *<* 7.2 delivery risk at 1-, 2-, and 4-hour forecast horizons. Event frequency is reported as test events/test patients (percentage). Expected-to-observed (E/O) ratio is mean predicted risk divided by observed event prevalence; values near 1 indicate agreement in the mean.

| Horizon (h) | Landmark (h) | Events | Raw E/O | Raw Brier | Recalibrated E/O | Recalibrated Brier |
|---|---|---|---|---|---|---|
| 1 | 1 | 9/3,405 (0.3%) | 10.55 | 0.0033 | 1.11 | 0.0031 |
|  | 6 | 24/2,886 (0.8%) | 2.65 | 0.0081 | 0.79 | 0.0083 |
|  | 12 | 43/1,907 (2.3%) | 1.19 | 0.0200 | 0.69 | 0.0198 |
|  | 20 | 17/853 (2.0%) | 1.46 | 0.0191 | 1.96 | 0.0196 |
| 2 | 1 | 19/3,405 (0.6%) | 6.89 | 0.0066 | 1.82 | 0.0068 |
|  | 6 | 42/2,886 (1.5%) | 2.74 | 0.0144 | 1.17 | 0.0146 |
|  | 12 | 84/1,907 (4.4%) | 1.37 | 0.0385 | 0.73 | 0.0378 |
|  | 20 | 33/853 (3.9%) | 1.86 | 0.0368 | 1.49 | 0.0370 |
| 4 | 1 | 47/3,405 (1.4%) | 4.48 | 0.0159 | 1.50 | 0.0142 |
|  | 6 | 94/2,886 (3.3%) | 2.32 | 0.0330 | 0.99 | 0.0311 |
|  | 12 | 151/1,907 (7.9%) | 1.23 | 0.0682 | 0.75 | 0.0677 |
|  | 20 | 70/853 (8.2%) | 1.50 | 0.0724 | 1.33 | 0.0721 |

**Figure K.2:**
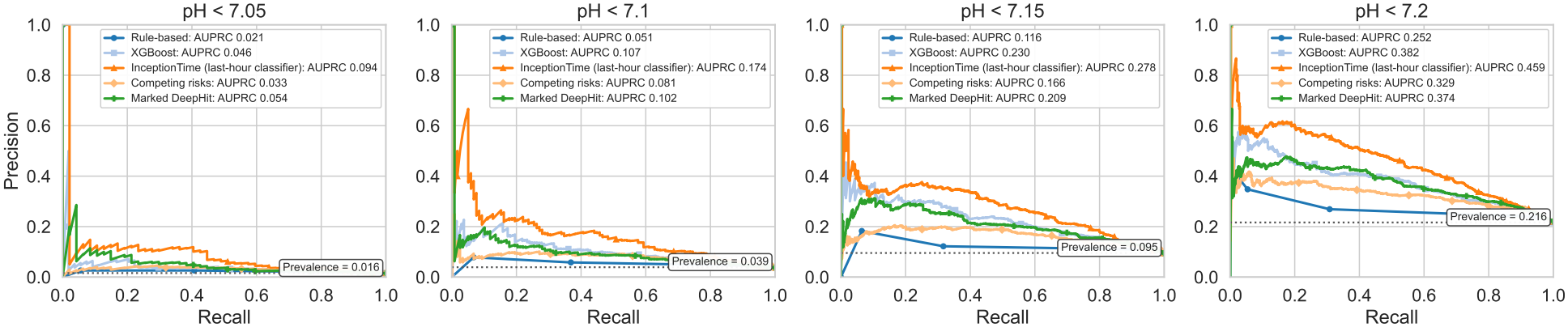
Held-out precision-recall curves at tracing end for acidemia classification at each cord-pH threshold. Average precision and the prevalence baseline are reported in the panel labels; AP lift for the distinct acidemic-delivery-within-one-hour endpoint is reported in Appendix G.

**Figure K.3:**
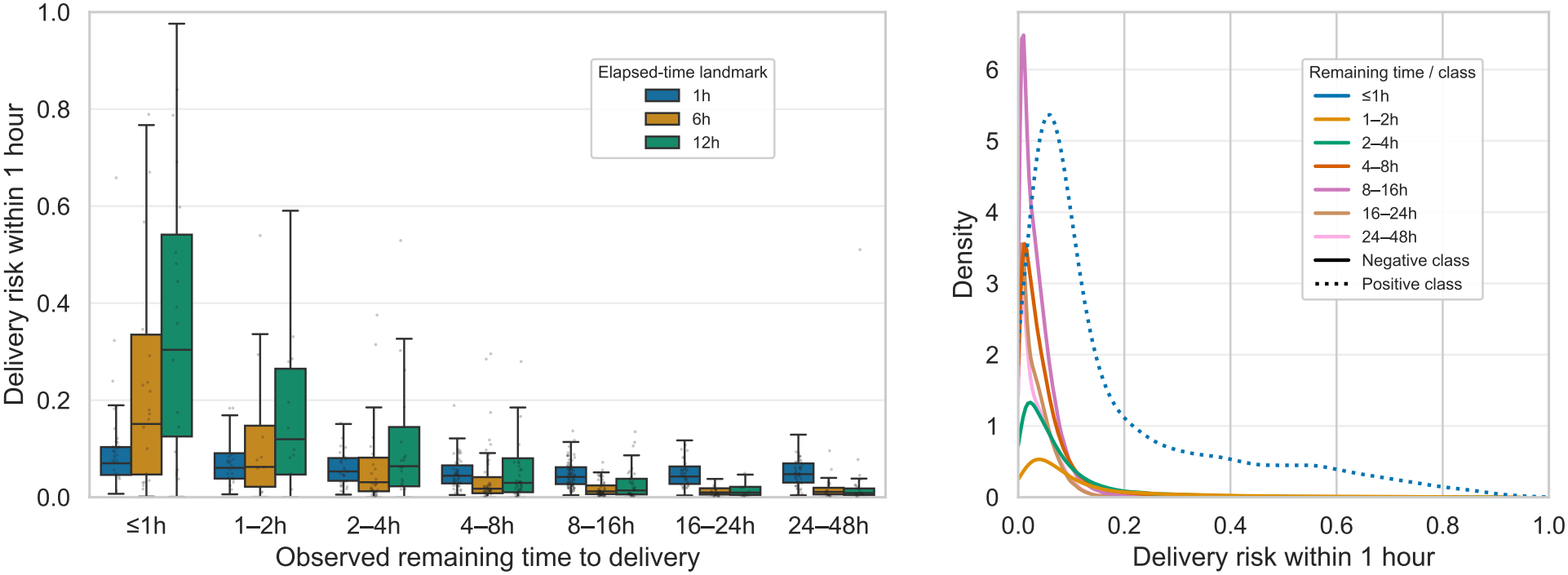
Marked DeepHit predictions for delivery within one hour. The left panel shows predicted risk by observed remaining-time band and elapsed-time landmark on a single 0–1 probability scale; the right panel shows risk distributions by remaining-time band.

## References

Z. Alfirevic, D. Devane, G. M. Gyte, and A. Cuthbert. Continuous cardiotocography (CTG) as a form of electronic fetal monitoring (EFM) for fetal assessment during labour. Cochrane Database Syst Rev, 2(2):Cd006066, February 2017. ISSN 1361-6137. doi: 10.1002/14651858.CD006066.pub3.

Per Kragh Andersen, Ronald B. Geskus, Theo de Witte, and Hein Putter. Competing risks in epidemiology: Possibilities and pitfalls. International Journal of Epidemiology, 41(3):861–870, 2012. doi: 10.1093/ije/dyr213.

Laura Antolini, Patrizia Boracchi, and Elia Biganzoli. A time-dependent discrimination index for survival data. Statistics in Medicine, 24(24):3927–3944, 2005. doi: 10.1002/sim.2427.

Diogo Ayres-de-Campos, Catherine Y. Spong, Edwin Chandraharan, and FIGO Intrapartum Fetal Monitoring Expert Consensus Panel. FIGO consensus guidelines on intrapartum fetal monitoring: Car-diotocography. International Journal of Gynecology & Obstetrics, 131(1):13–24, 2015. ISSN 1879-3479. doi: 10.1016/j.ijgo.2015.06.020.

I. Ben M’Barek, G. Jauvion, and P. F. Ceccaldi. Computerized cardiotocography analysis during labor - A state-of-the-art review. Acta Obstet Gynecol Scand, 102(2):130–137, February 2023. ISSN 0001-6349 (Print) 0001-6349. doi: 10.1111/aogs.14498.

Alison G. Cahill, Methodius G. Tuuli, Molly J. Stout, Julia D. López, and George A. Macones. A prospective cohort study of fetal heart rate monitoring: Deceleration area is predictive of fetal acidemia. American Journal of Obstetrics and Gynecology, 218(5):523.e1–523.e12, May 2018. ISSN 0002-9378. doi: 10.1016/j.ajog.2018.01.026.

M. Campanile, P. D’Alessandro, L. Della Corte, G. Saccone, S. Tagliaferri, B. Arduino, G. Esposito, F. G. Esposito, A. Raffone, M. G. Signorini, G. Magenes, M. Di Tommaso, S. Xodo, F. Zullo, and V. Berghella. Intrapartum cardiotocography with and without computer analysis: A systematic review and meta-analysis of randomized controlled trials. J Matern Fetal Neonatal Med, 33 (13):2284–2290, July 2020. ISSN 1476-4954. doi: 10.1080/14767058.2018.1542676.

Paolo Ivo Cavoretto, Anna Seidenari, and Antonio Farina. Hazard and cumulative incidence of umbilical cord metabolic acidemia at birth in fetuses experiencing the second stage of labor and pathologic intrapartum fetal heart rate requiring expedited delivery. Archives of Gynecology and Obstetrics, 307(4):1225–1232, 2023.

Václav Chudáček, JiříSpilka, Miroslav Burša, Petr Janků, Lukáš Hruban, Michal Huptych, and Lenka Lhotská. Open access intrapartum CTG database. BMC Pregnancy and Childbirth, 14:16, 2014. doi: 10.1186/1471-2393-14-16.

D. R. Cox. Regression models and life-tables. Journal of the Royal Statistical Society: Series B, 34 (2):187–202, 1972. doi: 10.1111/j.2517-6161.1972.tb00899.x.

Nan Du, Hanjun Dai, Rakshit Trivedi, Utkarsh Upadhyay, Manuel Gomez-Rodriguez, and Le Song. Recurrent marked temporal point processes: Embedding event history to vector. In Proceedings of the 22nd ACM SIGKDD International Conference on Knowledge Discovery and Data Mining, pages 1555–1564, 2016. doi: 10.1145/2939672.2939875.

Mark I. Evans, David W. Britt, Shara M. Evans, and Lawrence D. Devoe. Improving the interpretation of electronic fetal monitoring: The fetal reserve index. American Journal of Obstetrics and Gynecology, 228(5, Supplement):S1129–S1143, May 2023. ISSN 0002-9378. doi: 10.1016/j.ajog.2022.11.1275.

Helen Feltovich. Cervical evaluation. Obstetrics & Gynecology, 130(1):51–63, July 2017. doi: 10.1097/AOG.0000000000002106.

Jason P. Fine and Robert J. Gray. A proportional hazards model for the subdistribution of a competing risk. Journal of the American Statistical Association, 94(446):496–509, 1999. doi: 10.1080/01621459.1999.10474144.

Antoniya Georgieva, Patrice Abry, Václav Chudáček, Petar M. Djurić, Martin G. Frasch, René Kok Christopher A. Lear, Sebastiaan N. Lemmens, Inês Nunes, Aris T. Papageorghiou, et al. Computer-based intrapartum fetal monitoring and beyond: A review of the 2nd workshop on signal processing and monitoring in labor. Acta Obstetricia et Gynecologica Scandinavica, 98(9):1207–1217, 2019. doi: 10.1111/aogs.13639.

Hassan Ismail Fawaz, Germain Forestier, Jonathan Weber, Lhassane Idoumghar, and Pierre-Alain Muller. Deep learning for time series classification: A review. Data Mining and Knowledge Discovery, 33(4):917–963, 2019. doi: 10.1007/s10618-019-00619-1.

Hassan Ismail Fawaz, Benjamin Lucas, Germain Forestier, Charlotte Pelletier, Daniel F. Schmidt, Jonathan Weber, Geoffrey I. Webb, Lhassane Idoumghar, Pierre-Alain Muller, and François Petitjean. InceptionTime: Finding AlexNet for time series classification. Data Mining and Knowledge Discovery, 34(6):1936–1962, 2020. doi: 10.1007/s10618-020-00710-y.

Robert E. Kearney, Yvonne W. Wu, Johann Vargas-Calixto, Michael W. Kuzniewicz, Marie-Coralie Cornet, Heather Forquer, Lawrence Gerstley, Emily Hamilton, and Philip A. Warrick. Construction of a comprehensive fetal monitoring database for the study of perinatal hypoxic ischemic encephalopathy. Methods X, 12, June 2024. ISSN 2215-0161. doi: 10.1016/j.mex.2024.102664.

Håvard Kvamme, Ørnulf Borgan, and Ida Scheel. Time-to-event prediction with neural networks and Cox regression. Journal of Machine Learning Research, 20(129):1–30, 2019.

Changhee Lee, William Zame, Jinsung Yoon, and Mihaela van der Schaar. DeepHit: A Deep Learning Approach to Survival Analysis With Competing Risks. Proceedings of the AAAI Conference on Artificial Intelligence, 32(1), April 2018. ISSN 2374-3468. doi: 10.1609/aaai.v32i1.11842.

Ilya Loshchilov and Frank Hutter. SGDR: Stochastic gradient descent with warm restarts. In International Conference on Learning Representations, 2017. URL https://openreview.net/forum?id=Skq89Scxx.

G. A. Macones, G. D. Hankins, C. Y. Spong, J. Hauth, and T. Moore. The 2008 National Institute of Child Health and Human Development workshop report on electronic fetal monitoring: Update on definitions, interpretation, and research guidelines. Obstet Gynecol, 112(3):661–6, September 2008. ISSN 0029-7844 (Print) 0029-7844. doi: 10.1097/AOG.0b013e3181841395.

G. L. Malin, R. K. Morris, and K. S. Khan. Strength of association between umbilical cord pH and perinatal and long term outcomes: Systematic review and meta-analysis. BMJ (Clinical research ed.), 340:c1471, 2010. doi: 10.1136/bmj.c1471.

J. A. Martin, B. E. Hamilton, P. D. Sutton, S. J. Ventura, F. Menacker, and M. L. Munson. Births: Final data for 2002. Natl Vital Stat Rep, 52(10):1–113, December 2003. ISSN 1551-8922 (Print) 1551-8922.

Jennifer A. McCoy, Lisa D. Levine, Guangya Wan, Corey Chivers, Joseph Teel, and William G. La Cava. Intrapartum electronic fetal heart rate monitoring to predict acidemia at birth with the use of deep learning. American Journal of Obstetrics and Gynecology, April 2024. ISSN 0002-9378. doi: 10.1016/j.ajog.2024.04.022.

Lochana Mendis, Marimuthu Palaniswami, Fiona Brownfoot, and Emerson Keenan. Computerised Cardiotocography Analysis for the Automated Detection of Fetal Compromise during Labour: A Review. Bioengineering, 10(9):1007, September 2023. ISSN 2306-5354. doi: 10.3390/bioengineering10091007.

Lochana Mendis, Marimuthu Palaniswami, Emerson Keenan, and Fiona Brownfoot. Rapid detection of fetal compromise using input length invariant deep learning on fetal heart rate signals. Scientific Reports, 14(1):12615, June 2024. ISSN 2045-2322. doi: 10.1038/s41598-024-63108-6.

Lochana Mendis, Debjyoti Karmakar, Marimuthu Palaniswami, Fiona Brownfoot, and Emerson Keenan. Cross-Database Evaluation of Deep Learning Methods for Intrapartum Cardiotocogra-phy Classification. IEEE journal of translational engineering in health and medicine, 13:123–135, 2025. ISSN 2168-2372. doi: 10.1109/JTEHM.2025.3548401.

Karin B. Nelson, James M. Dambrosia, Tricia Y. Ting, and Judith K. Grether. Uncertain value of electronic fetal monitoring in predicting cerebral palsy. New England Journal of Medicine, 334(10):613–619, 1996. doi: 10.1056/NEJM199603073341001.

Petrozziello, I. Jordanov, T. Aris Papageorghiou, W. G. Christopher Redman, and A. Georgieva. Deep Learning for Continuous Electronic Fetal Monitoring in Labor. Annu Int Conf IEEE Eng Med Biol Soc, 2018:5866–5869, July 2018. ISSN 2375-7477. doi: 10.1109/embc.2018.8513625.

Alessio Petrozziello, Christopher W. G. Redman, Aris T. Papageorghiou, Ivan Jordanov, and Antoniya Georgieva. Multimodal Convolutional Neural Networks to Detect Fetal Compromise During Labor and Delivery. IEEE Access, 7:112026–112036, 2019. ISSN 2169-3536. doi: 10.1109/ACCESS.2019.2933368.

H. Putter, M. Fiocco, and R. B. Geskus. Tutorial in biostatistics: Competing risks and multi-state models. Statistics in Medicine, 26(11):2389–2430, 2007. doi: 10.1002/sim.2712.

Alex Reinhart. A review of self-exciting spatiotemporal point processes and their applications. Statistical Science, 33(3):299–318, 2018. doi: 10.1214/17-STS629.

Ayodeji Sanusi, Yuanfan Ye, Ashley N. Battarbee, Rachel Sinkey, Rebecca Pearlman, Danyon Beitel, Jeff M. Szychowski, Alan T. N. Tita, and Akila Subramaniam. Predicting spontaneous labor beyond 39 weeks among low-risk expectantly managed pregnant patients. American Journal of Perinatology, 40(16):1725–1731, May 2023. doi: 10.1055/a-2099-4395.

Oleksandr Shchur, Ali Caner Türkmen, Tim Januschowski, and Stephan Günnemann. Neural temporal point processes: A review. In Proceedings of the Thirtieth International Joint Conference on Artificial Intelligence (IJCAI), pages 4585–4593, 2021.

Jiri Spilka, Jordan Frécon, Roberto Leonarduzzi, Nelly Pustelnik, Patrice Abry, and Muriel Doret. Sparse support vector machine for intrapartum fetal heart rate classification. IEEE Journal of Biomedical and Health Informatics, 21(3):664–671, 2017. doi: 10.1109/JBHI.2016.2546312.

Ellen L. Tilden, Aaron B. Caughey, Mia Ahlberg, Louise Lundborg, Anna-Karin Wikström, Xingrong Liu, Kevin Ng, Jodi Lapidus, and Anna Sandström. Latent phase duration and associated outcomes: A contemporary, population-based observational study. American Journal of Obstetrics and Gynecology, 228(5):S1025–S1036.e9, May 2023. doi: 10.1016/j.ajog.2022.10.003.

Hans C. van Houwelingen. Dynamic prediction by landmarking in event history analysis. Scandinavian Journal of Statistics, 34(1):70–85, 2007. doi: 10.1111/j.1467-9469.2006.00529.x.

Johann Vargas-Calixto, Michael W Kuzniewicz, Marie-Coralie Cornet, Yvonne W Wu, Aditi Lahiri, Lawrence Gerstley, John Parker, Philip A Warrick, and Robert E Kearney. Time-Surrogate Variables Enhance the Association Between Cardiotocographic Features and Intrapartum Hypoxic-Ischemic Encephalopathy. PLOS Digital Health, 5 (8):e0001111, August 2026. ISSN 2767-3170. doi: 10.1371/journal.pdig.0001111. URL http://dx.doi.org/10.1371/journal.pdig.0001111.

Yingye Zheng and Patrick J. Heagerty. Partly conditional survival models for longitudinal data. Biometrics. Journal of the International Biometric Society, 61(2):379–391, 2005. doi: 10.1111/j.1541-0420.2005.00323.x.

